# GPT-4, an artificial intelligence large language model, exhibits high levels of accuracy on dermatology specialty certificate exam questions

**DOI:** 10.1101/2023.07.13.23292418

**Authors:** Meghna Shetty, Michael Ettlinger, Magnus Lynch

## Abstract

Artificial Intelligence (AI) has shown considerable potential within medical fields including dermatology. In recent years a new form of AI, large language models, has shown impressive performance in complex textual reasoning across a wide range of domains including standardised medical licensing exam questions. Here, we compare the performance of different models within the GPT family (GPT-3, GPT-3.5, and GPT-4) on 89 publicly available sample questions from the Dermatology specialty certificate examination. We find that despite no specific training on dermatological text, GPT-4, the most advanced large language model, exhibits remarkable accuracy - answering in excess of 85% of questions correctly, at a level that would likely be sufficient to pass the SCE exam.

---

Artificial Intelligence (AI) has shown considerable potential within a wide range of medical fields including dermatology ^1,2^. Recent years have seen the rapid advancement of “large language models”. These models are trained on very large quantities of textual data permitting complex reasoning to emerge. The best-performing models at the present time are the GPT family of models developed by openAI. In recent years GPT-3, GPT-3.5, and GPT-4 have been released each of which contains a larger number of parameters and better performance.

The largest, most advanced, large language model, GPT4, was released recently ^3^ and has shown remarkable performance across a wide range of domains including standardised general medical exam questions ^4^. For specialist areas within medicine such as dermatology, there is far less publicly available information, and it is unclear whether it will perform at such a high level. Here, we compare the performance of different models within the GPT family (GPT-3, GPT-3.5, and GPT-4) on 89 publicly available sample questions from the Dermatology specialty certificate (SCE) examination ^5^.

We downloaded the text of 89 questions from the MRCP UK Dermatology sample question dataset and their corresponding answers. The AI language model is “prompted” with arbitrary text and returns a textual answer in response. Using the OpenAI GPT application programming interface (API) developer preview we developed a script to automatically prompt the GPT API with the unedited full text of the multiple choice question followed immediately by the phrase “Only answer with the letter of the answer; do not elaborate further\n\nAnswer Letter:” This ensured the API only returned a letter per answer and not an explanation. We used the publicly available versions of the models and did not perform any fine-tuning training with medical or dermatological text. The answer was scored as correct where the letter response output by the AI matched the correct response. The API parameters were temperature=0, max_tokens=1, top_p=1, frequency_penalty=0.0, presence_penalty=0.0. For GPT-3, we used the model text-davinci-003; for GPT-3.5, we used gpt-3.5-turbo, and for GPT-4, we used “gpt-4.”. Text from tables in the questions was included in the prompt but images were not included.

We first compared the relative performance of the different large language models (Table 1). GPT-3 answered 43 questions correctly (48.31%), GPT3.5 answered 54 questions correctly (60.57%), and GPT-4 outperformed both by answering 76 questions correctly (85.39%).

**Table 1:** Comparison of responses of versions of Chat GPT 3 versus 3.5 versus 4 in its performance in Dermatology SCE questions:

|  | Question type | GPT 3 | GPT 3.5 | GPT 4 |
| --- | --- | --- | --- | --- |
| Total Questions | All questions | 89 | 89 | 89 |
| Correct answers |  | 43 | 54 | 76 |
| Percentage correct |  | 48.31% | 60.67% | 85.39% |
| Total Questions | Questions with images | 4 | 4 | 4 |
| Correct answers |  | 2 | 3 | 3 |
| Percentage correct |  | 50.00% | 75.00% | 75.00% |
| Total Questions | Questions with tables | 6 | 6 | 6 |
| Correct answers |  | 0 | 3 | 4 |
| Percentage correct |  | 0.00% | 50.00% | 66.67% |
| Total Questions | Questions without tables or images | 79 | 79 | 79 |
| Correct answers |  | 41 | 48 | 69 |
| Percentage correct |  | 51.90% | 60.76% | 87.34% |

GPT4 accurately answered 75% of the image questions and 66.67% of the table questions among the ten questions that included an image or table. For the remaining 79 questions without images or tables, GPT-4 achieved an impressive 87.34% accuracy (Table 1). To examine reasoning capability in more detail, we executed the script again with the question and the prompt “Elaborate on your answer:” for both correct and incorrect responses (www.github.com/thelynchlab/gpt4). These results highlight the incredible advances that large language models have made in answering questions relating to specialist dermatological conditions.

Images in the questions were not included in the information supplied to the model, yet despite this 75% of the image questions were answered correctly, however, GPT-4 is capable of reading and interpreting tables. When we examined the questions that were incorrectly answered, we observed that GPT often provided the correct diagnosis but did not select the most appropriate treatment options. For example, GPT-4 correctly made the diagnosis of pyoderma gangrenosum but suggested dapsone as the appropriate initial intervention, whereas the correct answer is prednisolone.

In summary, with no specific training for medical or dermatology questions, GPT-4 exhibited remarkable accuracy in answering text-based dermatology multiple-choice scenarios at a level that would likely be sufficient to pass the SCE exam. In future studies, it will be important to assess the performance of the model on more realistic clinical scenarios in combination with medical images. It will also be of value to assess whether “fine-tuning” the model with clinical images and non-publicly available text relating to dermatological diagnosis leads to superior performance, particularly for rarer conditions.

Large language models, particularly in combination with clinical image processing have the potential to improve the provision of dermatological services. Obvious applications would include the descriptive interpretation of dermoscopy and dermatopathology images and aid in the diagnosis of rare conditions. These capabilities are likely to be of particular value in resource poor environments where access to highly experienced specialists is limited.

## Data Availability

All data produced are available online at
https://www.github.com/thelynchlab/gpt4

## Acknowledgments

We would like to thank Open AI for allowing us to use its resources in the field of research.

